# Bidirectional associations between COVID-19 and psychiatric disorder: a study of 62,354 COVID-19 cases

**DOI:** 10.1101/2020.08.14.20175190

**Authors:** Maxime Taquet, Sierra Luciano, John R Geddes, Paul J Harrison

## Abstract

**Background:** Adverse mental health consequences of COVID-19, including anxiety and depression, have been widely predicted but not yet accurately measured. There are a range of physical health risk factors for COVID-19, but it is not known if there are also psychiatric risk factors.

**Methods:** We addressed both questions using cohort studies derived from an electronic health records (EHR) network of 69 million patients including over 62,000 cases of COVID-19. Propensity score matching was used to control for confounding by risk factors for COVID-19 and for more severe illness.

**Findings:** In patients with no prior psychiatric history, COVID-19 was associated with an increased incidence of psychiatric diagnoses in the three months after infection compared to 6 other health events (hazard ratio [95% CI] 2.1 [1.8–2.5] compared to influenza; 1.7 [1.5–1.9] compared to other respiratory tract infections; 1.6 [1.4–1.9] compared to skin infection; 1.6 [1.3–1.9] compared to cholelithiasis; 2.2 [1.9–2.6] compared to urolithiasis, and 2.1 [1.9–2.5] compared to fracture of a large bone; all p< 0.0001). The increase was greatest for anxiety disorders but also present for depression, insomnia, and dementia. The results were robust to several sensitivity analyses. There was a ∼30% reduction in psychiatric diagnoses in the total EHR population over the same period. A psychiatric diagnosis in the previous year was associated with a 65% higher incidence of COVID-19 (relative risk 1.65, 95% CI: 1.59–1.71, p< 0.0001). This was independent of known physical health risk factors for COVID-19.

**Interpretation:** COVID-19 infection has both psychiatric sequelae and psychiatric antecedents. Survivors have an increased rate of new onset psychiatric disorders, and prior psychiatric disorders are associated with a higher risk of COVID-19. The findings have implications for research into aetiology and highlight the need for clinical services to provide multidisciplinary follow-up, and prompt detection and treatment.

## Introduction

From the early stages of the coronavirus disease 2019 (COVID-19) pandemic, concerns have been raised about its impact on mental health^1–3^ and on patients with mental illness^4^. Yet a few months later, we still know little about the mental health consequences of COVID-19 (its psychiatric sequelae) and the susceptibility of patients with mental illness to COVID-19 (its psychiatric antecedents).

Several surveys have suggested that patients with COVID-19 experience symptoms of anxiety^5–9^ (including post-traumatic stress disorder after discharge^10^), depression^5–9,11,12^ and insomnia^9^. CORONERVE, a UK-wide surveillance program has identified 23 patients with a psychiatric diagnosis following infection with SARS-CoV-2^13^. A meta-analysis pooled data from studies estimating the incidence of psychiatric disorders following the severe acute respiratory syndrome (SARS) and Middle East respiratory syndrome (MERS), suggesting that coronavirus infections may lead to delirium, anxiety, depression, manic symptoms, poor memory, and insomnia^14^. However cohort studies of patients with COVID-19 with adequate control and follow-up are urgently needed to quantify the incidence of psychiatric sequelae following infection with the virus.

A separate question is whether pre-existing psychiatric disorder affects susceptibility to COVID-19 infection. A study based on 908 cases from UK Biobank revealed that self-reported symptoms of mental illness were associated with an increased risk of hospitalisation for COVID-19 but these were not robust to adjustment for several confounders^15^. A case-control study based on the electronic health records (EHR) of 843 patients with COVID-19 found that depression, anxiety, and dementia might increase the odds of being diagnosed with COVID-19^16^. However, reliable estimation of possible increased risk of COVID-19 among patients with a psychiatric illness requires large, well-controlled cohort studies.

In this EHR network cohort study, we assessed the psychiatric sequelae and antecedents of COVID-19 using data from 69 million individuals, 62,354 of whom have had a diagnosis of COVID-19.

## Methods

### Data and study design

We used data from TriNetX Analytics Network (http://www.trinetx.com), a global federated research network capturing anonymized data from EHR in 54 healthcare organizations in the USA, totalling 69.8 million patients. The TriNetX platform and its functionalities have been described elsewhere^17^. The healthcare organizations are a mixture of hospitals, primary care, and specialist providers. TriNetX has a waiver from the Western Institutional Review Board since only de-identified summary statistics are provided. Available data include demographics, diagnoses (using ICD-10 codes), procedures, and measurements (e.g. lab results, body mass index). Most captured data were from 2007 onwards, and data are updated on average every 24 days.

Using the TriNetX user interface, cohorts can be created based on specified inclusion and exclusion criteria and compared for outcomes of interest over specified time periods. Cohorts were matched for confounding variables using the built-in propensity score matching capability. Outcomes of interest were then compared between matched cohorts. This study followed the STROBE reporting guidelines.

### Variables of interest and their coding

We defined a diagnosis of COVID-19 as one of the following diagnoses, recorded on or after January 20, 2020 (date of the first recorded COVID-19 case in the USA): COVID-19 (U07.1 and U07.2); Pneumonia due to SARS-associated coronavirus (J12.81); Other coronavirus as the cause of disease classified elsewhere (B97.29); or Coronavirus infection unspecified (B34.2). Inclusion of the latter three definitions (which make up 7.3% of the total COVID-19 sample) were included to capture the early stage of the pandemic when the ICD code for COVID-19 (U07) was not yet defined. We define a psychiatric illness as any of the ICD-10 codes F20-F48 corresponding to psychotic (F20-F29), mood (F30-F39), and anxiety (F40-F48) disorders.

We identified a set of established and suspected risk factors for COVID-19^18–20^: age, sex, race, obesity, hypertension, diabetes, chronic kidney disease, asthma, chronic lower respiratory diseases, nicotine dependence, ischaemic heart disease, and other forms of heart disease. To capture these risk factors in patients’ EHR, we used 28 variables (e.g. diabetes was separated into Type 1 and Type 2, hypertension was represented both as a diagnosis and as a measurement of systolic and diastolic blood pressure, etc.). We also identified an additional set of established risk factors for death due to COVID-19^21^ (which we take to be risk factors for severe illness): cancer (and haematological cancer in particular), chronic liver disease, stroke, dementia, organ transplant, rheumatoid arthritis, lupus, psoriasis, and other immunosuppression. These risk factors were captured using 22 variables from patients’ EHR.

More details about these variables are provided in the appendix, pp. 1–2.

### Analysis of psychiatric sequelae

To assess the psychiatric sequelae of COVID-19 and compare them to other acute health events, we produced matched cohorts of patients who had been diagnosed with another health event. The other health events were selected to represent a wide range of common acute presentations (some clinically similar to COVID-19 and others very different). These control health events comprised: (i) influenza, (ii) another respiratory tract infection, (iii) skin infection, (iv) cholelithiasis, (v) urolithiasis, and (vi) fracture of a large bone (see appendix, p. 2).

All seven cohorts (COVID-19 and six control health events) included all patients over the age of ten who had the corresponding health event on or after January 20, 2020. We excluded patients who had a psychiatric diagnosis recorded at any time before January 20, 2020, and patients who had died by the time of the analysis (August 1, 2020). Cohorts were matched (details shown below) for the 50 variables mentioned above: the 28 variables capturing risk factors for COVID-19 and the 22 variables capturing risk factors for more severe COVID-19 illness.

The primary outcome was the incidence of a first psychiatric diagnosis (F20-F48) over a period from 14 days to 90 days after a diagnosis of COVID-19 represented by a hazard ratio (HR) and by the estimated probability of outcome over that period. Other outcomes included incidence of subcategories of these psychiatric illnesses. We also assessed for dementia and insomnia as they are thought to be potential sequelae of COVID-19^9,14^. For dementia, the analysis was repeated among patients over the age of 65 years.

Besides using six different control cohorts, the robustness of the findings was tested by repeating the analysis (i) after excluding individuals whose race was unknown (in case this differentially affected cohorts), (ii) by restricting the diagnosis of COVID-19 to confirmed diagnoses (ICD-10 code U07.1), and (iii) by focusing on patients who made at least one healthcare visit between 14 and 90 days after the health event (in case of differential drop-out rates between cohorts).

Besides the explanation that COVID-19 itself leads to increased rates of psychiatric sequelae, we tested three alternative hypotheses which could explain differences in outcomes between cohorts. The “severity” hypothesis posits that differences in rates of psychiatric sequelae are due to differences in the severity of the health event (e.g. COVID-19 might lead to more severe presentations than influenza). We tested this hypothesis by limiting the cohorts to patients with the least severe presentations (taken to be those not requiring inpatient admission). If the hypothesis were correct, the difference in rates of psychiatric sequelae between these cohorts would be substantially smaller than in the original cohorts. The “overall incidence” hypothesis posits that, compared to control health events, COVID-19 was often diagnosed at a time when more people (regardless of their COVID-19 status) were diagnosed with a psychiatric illness (e.g. due to lockdown, financial strain, seasonality, etc.). It would follow that the increased rate of psychiatric sequelae following COVID-19 simply reflects a change in incidence of psychiatric illness in the general population. We tested this hypothesis by comparing the evolution of the incidence of psychiatric diagnoses, COVID-19, and control health events across the study period. Finally, the “contextual factors” hypothesis posits that COVID-19 was mostly diagnosed at a time when having *any* health event would have increased the risk of psychiatric sequelae (e.g. because of overwhelmed health services, fear of COVID-19, limited social support, etc.). Assuming that contextual factors may have changed substantially between January and April 2020, we tested this hypothesis by comparing the rate of psychiatric sequelae of health events before vs. after April 1, 2020 and by comparing the rate of psychiatric sequelae between COVID-19 and control health events after April 1, 2020.

Further details on the outcome definitions and sensitivity analyses are provided in the appendix (pp. 2–5).

### Analysis of psychiatric antecedents

We tested whether patients with a recent diagnosis of psychiatric illness were at a higher risk of developing COVID-19 compared to a matched cohort of patients with otherwise similar risk factors for COVID-19.

Two cohorts were defined. The first cohort included all patients over the age of 18 who had a diagnosis of a psychiatric illness recorded in their EHR in the past year (from January 21, 2019 to January 20, 2020). The second cohort had no psychiatric illness recorded in their EHR but did make a healthcare visit in the same period (thus excluding patients who made no contact with the participating healthcare organizations). We also defined separate cohorts for the three main classes of psychiatric illness (psychotic disorder [F20-F29], mood disorders [F30-F39], and anxiety disorders [F40-F48]). Patients who had died before January 20, 2020 were excluded from both cohorts.

Cohorts were matched for the 28 variables capturing risk factors for COVID-19 (see above). The primary outcome was the relative risk (RR) of being diagnosed with COVID-19 between matched cohorts. The robustness of the findings was tested by repeating the analysis in 5 scenarios: (i) limiting the cohorts to those with none of the physical risk factors for COVID-19, (ii) extending the window for a psychiatric diagnosis from one to three years before January 20, 2020 (iii) limiting the cohort to patients with a first diagnosis of psychiatric illness (i.e. with no diagnosis present before January 21, 2019), (iv) excluding patients with unknown race, and (v) redefining the primary outcome as a confirmed COVID-19 diagnosis.

More details on the sensitivity analyses are provided in the appendix pp. 3–5.

### Statistical analyses

Propensity score 1:1 matching used a greedy nearest neighbour matching with a caliper distance of 0.1 pooled standard deviations of the logit of the propensity score^22^. We considered any variable which had a standardized mean difference between cohorts lower than 0.1 to be well matched^23^. For the analysis of psychiatric sequelae, propensity score matching was directly applied to each cohort pair. For the analysis of psychiatric antecedents, given their much larger sample sizes (which exceeded the maximum number of 1.5 million patients possible per matched cohort), cohorts were first stratified by sex and age (from 18 years old to 30, 31 to 45, 46 to 60, 61 to 75, and 76 and over) and propensity score matching (including for age) was achieved within each stratum separately.

In the analysis of psychiatric sequelae, Kaplan-Meier analysis was conducted to estimate the probability of outcomes from 14 to 90 days. Comparisons between cohorts were made using a log-rank test. The HR was calculated using a proportional hazard model (with the survival package 3.2.3 in R) wherein the cohort to which the patient belonged was used as the independent variable. The proportional hazard assumption was tested using the generalized Schoenfeld approach^24^. If the assumption was violated, a piecewise constant HR was estimated by calculating a separate HR for the early and late phases of the follow-up period and the assumption was tested again in each sub-period (see appendix p. 5).

In the analysis of the psychiatric antecedents, the RR of being diagnosed with COVID-19 were calculated for each stratum and for the whole cohort. The null hypothesis that the outcome rate is equal in the two cohorts was tested using a χ^2^-test. A logistic regression was used to test for a potential association between age and RR (see appendix p. 6).

Statistical analyses were conducted in R version 3.4.3 except for the logrank tests which were performed within TriNetX. Statistical significance was set at two-sided p-values < 0.05.

## Results

### Characteristics of the cohorts and descriptive statistics

A total of 62,354 patients had a diagnosis of COVID-19 (Table 1). For the analysis of psychiatric sequelae, a subset of 44,779 patients who had no prior psychiatric illness and who had not died was used as the COVID-19 cohort. Successful matching was achieved between this cohort and cohorts with other acute health events (appendix pp. 7-18). For the analysis of psychiatric antecedents, a cohort of 1,729,837 patients with a psychiatric diagnosis between January 21, 2019 and January 20, 2020 was defined and successfully matched to a cohort of 1,729,837 patients who never had a psychiatric diagnosis (appendix p. 19).

**Table 1.** Baseline characteristics of the patients with a COVID-19 diagnosis

|  | n (%), mean (SD) |
| --- | --- |
| **COVID-19** |  |
| Diagnosis of COVID-19 | 62,354 (100.0) |
| of which confirmed diagnosis | 57,821 (92.7) |
| **DEMOGRAPHICS** |  |
| Age, mean (SD), y | 49.3 (19.7) |
| **Sex** |  |
| Female | 34,461 (55.3) |
| Male | 28,399 (45.5) |
| Other | 261 (0.4) |
| **Race** |  |
| White | 31,789 (51.0) |
| Black or African American | 14,700 (23.6) |
| Asian | 1,554 (2.5) |
| American Indian or Alaska Native | 329 (0.5) |
| Native Hawaiian or Other Pacific Islander | 107 (0.2) |
| Unknown | 13,875 (22.3) |
| **COMORBIDITIES** |  |
| **Obesity** |  |
| Overweight and obesity | 12,249 (19.6) |
| Body mass index, n with data (%) \| mean (SD), kg/m<sup>2</sup> | 23,728 (38.1) \| 28.1 (8.2) |
| **Hypertension** |  |
| Systolic Blood Pressure, n with data (%) \| mean (SD), mmHg | 41,011 (65.8) \| 128 (20.6) |
| Diastolic Blood Pressure, n with data (%) \| mean (SD), mmHg | 41,009 (65.8) \| 76.9 (13.1) |
| Hypertensive diseases | 21,228 (34.0) |
| **Diabetes mellitus** |  |
| Type 1 diabetes mellitus | 1,535 (2.5) |
| Type 2 diabetes mellitus | 10,998 (17.6) |
| **Chronic lower respiratory diseases** |  |
| Bronchitis; not specified as acute or chronic | 3,125 (5.0) |
| Simple and mucopurulent chronic bronchitis | 329 (0.5) |
| Unspecified chronic bronchitis | 388 (0.6) |
| Emphysema | 1,211 (1.9) |
| Other chronic obstructive pulmonary disease | 3,582 (5.7) |
| Asthma | 7,101 (11.4) |
| Bronchiectasis | 384 (0.6) |
| **Nicotine dependence** | 4,579 (7.3) |
| **Heart diseases** |  |
| Ischemic heart diseases | 6,579 (10.6) |
| Other forms of heart disease | 12,633 (20.3) |
| **Chronic kidney diseases** |  |
| Chronic kidney disease (CKD) | 5,554 (8.9) |
| Hypertensive chronic kidney disease | 2,890 (4.6) |
| **Chronic liver diseases** |  |
| Alcoholic liver disease | 351 (0.6) |
| Hepatic failure; not elsewhere classified | 502 (0.8) |
| Chronic hepatitis; not elsewhere classified | 83 (0.1) |
| Fibrosis and cirrhosis of liver | 775 (1.2) |
| Fatty (change of) liver; not elsewhere classified | 2,152 (3.5) |
| Chronic passive congestion of liver | 388 (0.6) |
| Portal hypertension | 322 (0.5) |
| Other specified diseases of liver | 1,502 (2.4) |
| **Cerebral infarction** | 1,910 (3.1) |
| **Dementia** |  |
| Vascular dementia | 558 (0.9) |
| Dementia in other diseases classified elsewhere | 740 (1.2) |
| Unspecified dementia | 1,794 (2.9) |
| Alzheimer's disease | 672 (1.1) |
| **Neoplasms** |  |
| Neoplasms | 12,655 (20.3) |
| Malignant neoplasms of lymphoid; hematopoietic and related tissue | 836 (1.3) |
| **Organ transplant** |  |
| Renal Transplantation Procedures | 137 (0.2) |
| Liver Transplantation Procedures | 44 (0.1) |
| **Psoriasis** | 669 (1.1) |
| **Rheumatoid arthritis** |  |
| Rheumatoid arthritis with rheumatoid factor | 301 (0.5) |
| Other rheumatoid arthritis | 982 (1.6) |
| **Systemic lupus erythematosus (SLE)** | 414 (0.7) |
| **Disorders involving the immune mechanism** | 1,532 (2.5) |
| **PSYCHIATRIC DIAGNOSES** |  |
| Psychiatric illness (F20-F48) | 15,980 (25.6) |
| Psychotic disorders (F20-29) | 1,219 (2.0) |
| Mood disorders (F30-F39) | 9,921 (15.9) |
| Anxiety disorder (F40-F48) | 12,145 (19.5) |
| **ADMISSIONS** |  |
| Critical care admission | 3490 (5.6) |
| Inpatient admission | 13152 (21.1) |

### Psychiatric sequelae of COVID-19

The estimated probabilities of psychiatric sequelae during the first 14-90 days following COVID-19 and other control health events are presented in Table 2 (the corresponding HR are reported in the appendix, p. 20). Compared to all six control health events, a diagnosis of COVID-19 led to significantly more first diagnoses of psychiatric illness (HR between 1.58 and 2.24, all p-values < 0.0001; Fig. 1 and appendix p. 21). At 90 days, the estimated probability of having been diagnosed with a new onset psychiatric illness following COVID-19 was 5.8% (95% CI: 5.2–6.4). The proportional hazard assumption was valid for three out of six control health events (influenza, other respiratory tract infection, and urolithiasis). For the other three events (skin infection, cholelithiasis, and fracture), there was evidence of non-proportionality and the HR tended to increase over time (appendix p. 22). However, the HR remained significantly greater than 1 for both the early and late phases of the follow-up period (all p< 0.0001, except for cholelithiasis in the early phase: p=0.0044). The most frequent psychiatric diagnosis following COVID-19 was anxiety disorder (HRs 1.59–2.62, all p-values < 0.0001) with a probability of outcome within 90 days of 4.7% (95% CI: 4.2–5.3). Among the anxiety disorders, adjustment disorder, generalized anxiety disorder, and to a lesser extent post-traumatic stress disorder and panic disorder were the most frequent (appendix pp. 23–24).

**Fig. 1.**
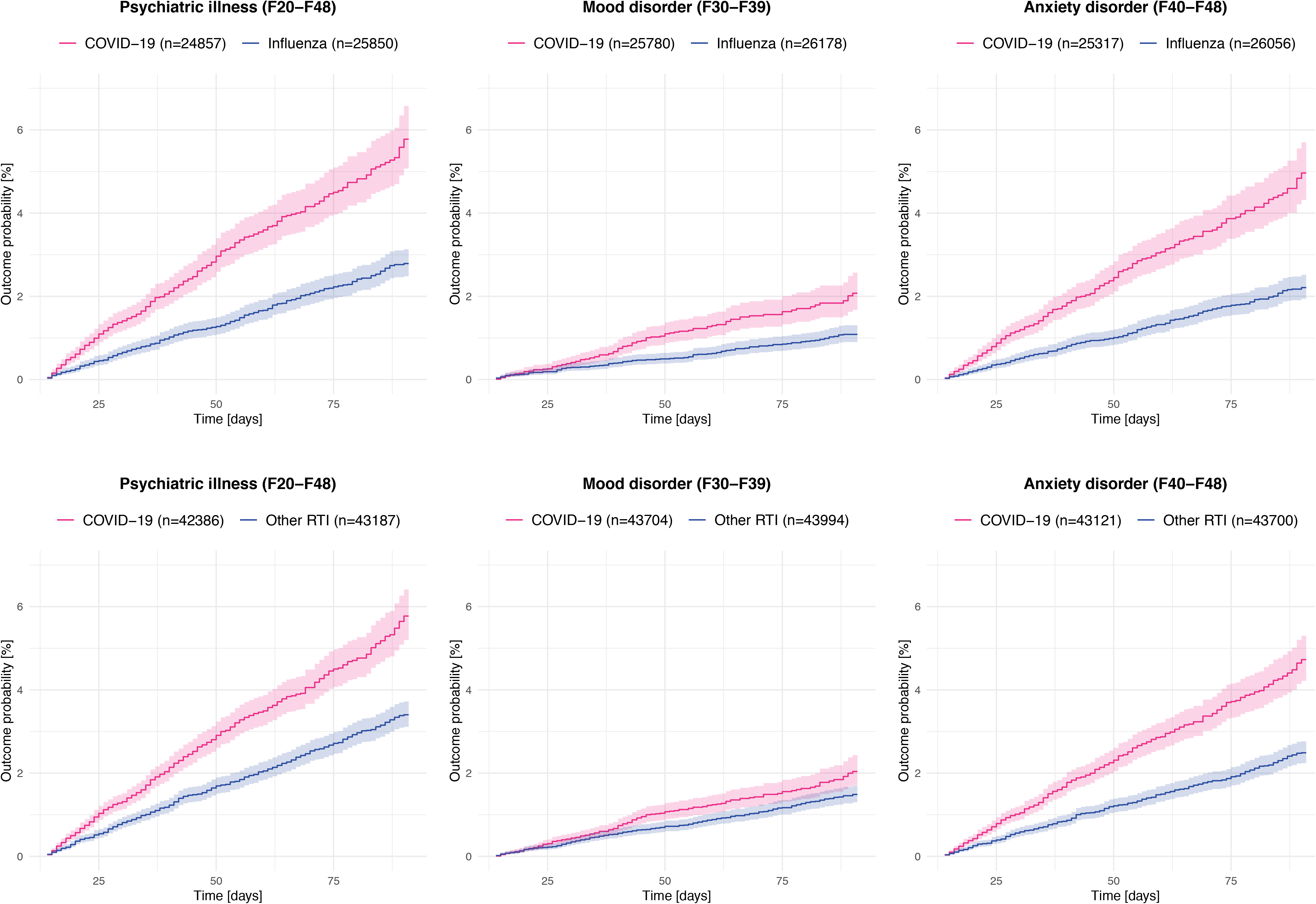
Kaplan-Meier curves representing the psychiatric sequelae of COVID-19 compared to influenza and other respiratory tract infections (RTI) The same curves for the other control health events are presented in the appendix, p. 21. Shaded areas represent 95% confidence intervals. The number of subjects within each cohort corresponds to all those that did not have the outcome before the follow-up period.

**Table 2.** Estimated incidence of first psychiatric diagnoses for the period 14–90 days after a diagnosis of COVID-19 compared to other health events.

| n in each matched cohort | COVID-19 | Influenza |  | Other RTI |  | Skin infection |  |
| --- | --- | --- | --- | --- | --- | --- | --- |
|  | - | 26497 |  | 44775 |  | 38977 |  |
|  | % (95% CI) | % (95% CI) | p | % (95% CI) | p | % (95% CI) | p |
| **Psychiatric illness** | 5.8 (5.2-6.4) | 2.8 (2.5-3.1) | <0.0001 | 3.4 (3.1-3.7) | <0.0001 | 3.3 (3-3.7) | <0.0001 |
| Psychotic disorder | 0.1 (0.08-0.2) | 0.04 (0.01-0.10) | 0.019 | 0.1 (0.06-0.16) | 0.23 | 0.15 (0.096-0.24) | 0.83 |
| Mood disorder | 2 (1.7-2.4) | 1.1 (0.9-1.3) | <0.0001 | 1.5 (1.3-1.7) | 0.0054 | 1.7 (1.5-1.9) | 0.55 |
| Anxiety disorder | 4.7 (4.2-5.3) | 2.2 (1.9-2.5) | <0.0001 | 2.5 (2.2-2.8) | <0.0001 | 2.4 (2.1-2.7) | <0.0001 |
| **Insomnia** | 1.9 (1.6-2.2) | 0.6 (0.5-0.8) | <0.0001 | 0.8 (0.7-1.0) | <0.0001 | 0.89 (0.73-1.1) | <0.0001 |
| **Dementia** | 0.44 (0.33-0.60) | 0.11 (0.06-0.20) | 0.00044 | 0.25 (0.18-0.35) | 0.00063 | 0.28 (0.20-0.39) | 0.13 |
| **Dementia (among 65+)** | 1.6 (1.2-2.1) | 0.66 (0.41-1.1) | 0.0043 | 0.84 (0.61-1.1) | 0.00071 | 0.70 (0.49-1.0) | 0.00069 |

| n in each matched cohort | Cholelithiasis |  | Urolithiasis |  | Fracture |  |
| --- | --- | --- | --- | --- | --- | --- |
|  | 19733 |  | 28827 |  | 37841 |  |
|  | % (95% CI) | p | % (95% CI) | p | % (95% CI) | p |
| **Psychiatric illness** | 3.2 (2.8-3.7) | <0.0001 | 2.5 (2.2-2.8) | <0.0001 | 2.5 (2.2-2.7) | <0.0001 |
| Psychotic disorder | 0.11 (0.054-0.24) | 0.21 | 0.044 (0.016-0.12) | 0.0051 | 0.16 (0.11-0.24) | 0.77 |
| Mood disorder | 1.6 (1.3-1.9) | 0.14 | 1.2 (1-1.4) | 0.00011 | 1.4 (1.2-1.6) | 0.005 |
| Anxiety disorder | 2.6 (2.2-3) | <0.0001 | 1.8 (1.6-2.1) | <0.0001 | 1.6 (1.4-1.8) | <0.0001 |
| **Insomnia** | 1.1 (0.88-1.4) | <0.0001 | 0.57 (0.43-0.74) | <0.0001 | 0.7 (0.57-0.85) | <0.0001 |
| **Dementia** | 0.24 (0.14-0.38) | <0.0001 | 0.16 (0.09-0.28) | <0.0001 | 0.34 (0.25-0.44) | 0.14 |
| **Dementia (among 65+)** | 0.58 (0.36-0.94) | <0.0001 | 0.60 (0.38-0.95) | <0.0001 | 0.94 (0.68-1.3) | 0.0036 |

P-values are obtained using a logrank test. A breakdown of the results for different diagnoses of the anxiety disorders and mood disorders categories is provided in Table 10 and 11 respectively (appendix pp. 24-25).

The probability of a first diagnosis of mood disorder within 14-90 days after COVID-19 was 2% (95% CI: 1.7–2.4). The corresponding hazard rate was comparable to that following a diagnosis of skin infection (HR 1.07, 95% CI: 0.87–1.31, p=0.55) or cholelithiasis (HR 1.22, 95% CI: 0.93–1.59, p = 0.14) but was significantly higher than the hazard rate hazard rate following a diagnosis of influenza (HR 1.79, 95% CI: 1.p=0.37, 2.33, p< 0.0001), another respiratory tract infection (HR 1.33, 95% CI: 1.09–1.63, p = 0.0054), urolithiasis (HR 1.62, 95% CI: 1.26–2.07, p=0.00011), or a fracture (HR.35, 95% CI: 1.094–1.67, p=0.005). Depressive episode was the most common first diagnosis of mood disorder (1.7%, 95% CI: 1.4–2.1; appendix p. 25).

There was a low probability of being newly diagnosed with a psychotic disorder in the 14-90 days following COVID-19 (0.1%, 95% CI: 0.08–0.2), broadly similar to the probability following control health events (Table 2). The probability of a first diagnosis of insomnia in the 90 days following COVID-19 was 1.9% (95% CI: 1.6–2.2) and insomnia was diagnosed significantly more than after control health events (HRs 1.85–3.29, all p-values < 0.0001). About 60% of the insomnia diagnoses were not accompanied by a concurrent diagnosis of an anxiety disorder (appendix p. 25). The probability of developing dementia was increased following a diagnosis of COVID-19 compared to all control health events (Table 2); among patients over the age of 65 years the risk was 1.6% (95% CI: 1.2–2.1), with HR between 1.89 and 3.18 (p-values in Table 2).

The increased risk of psychiatric sequelae after COVID-19 remained unchanged when the cohorts were limited to patients with known race (HR between 1.52 and 2.19, all p < 0.0001, appendix p. 26), patients with confirmed COVID-19 (HR between 1.63 and 2.28, all p < 0.0001, appendix p. 27), and patients who made at least one healthcare visit between 14 and 90 days after their health event (HR between 1.66 and 1.77, all p< 0.0001, appendix p. 28).

The elevated risk of psychiatric sequelae after COVID-19 compared to control health events could not be readily explained by differences in illness severity. Patients with COVID-19 requiring inpatient admission were more at risk of psychiatric sequelae than patients not needing an admission (HR 1.40, 95% CI: 1.06–1.85, p=0.019). However, when limiting cohorts to those not requiring inpatient admission, large differences in psychiatric sequelae remained between COVID-19 and the other cohorts (HR 1.54–2.23, all p< 0.0001, appendix p. 29).

Neither could the finding of higher rates of psychiatric disorder following COVID-19 be explained by a general increase in incidence of psychiatric diagnoses in the general population. Indeed diagnostic rates decreased from January to July 2020 (alongside a reduction in control health events), in contrast to the incidence of COVID-19 (Fig. 2 and appendix p. 30). In other words, the psychiatric sequelae of COVID-19 occurred against a backdrop of fewer psychiatric diagnoses being made in the total population.

**Fig. 2.**
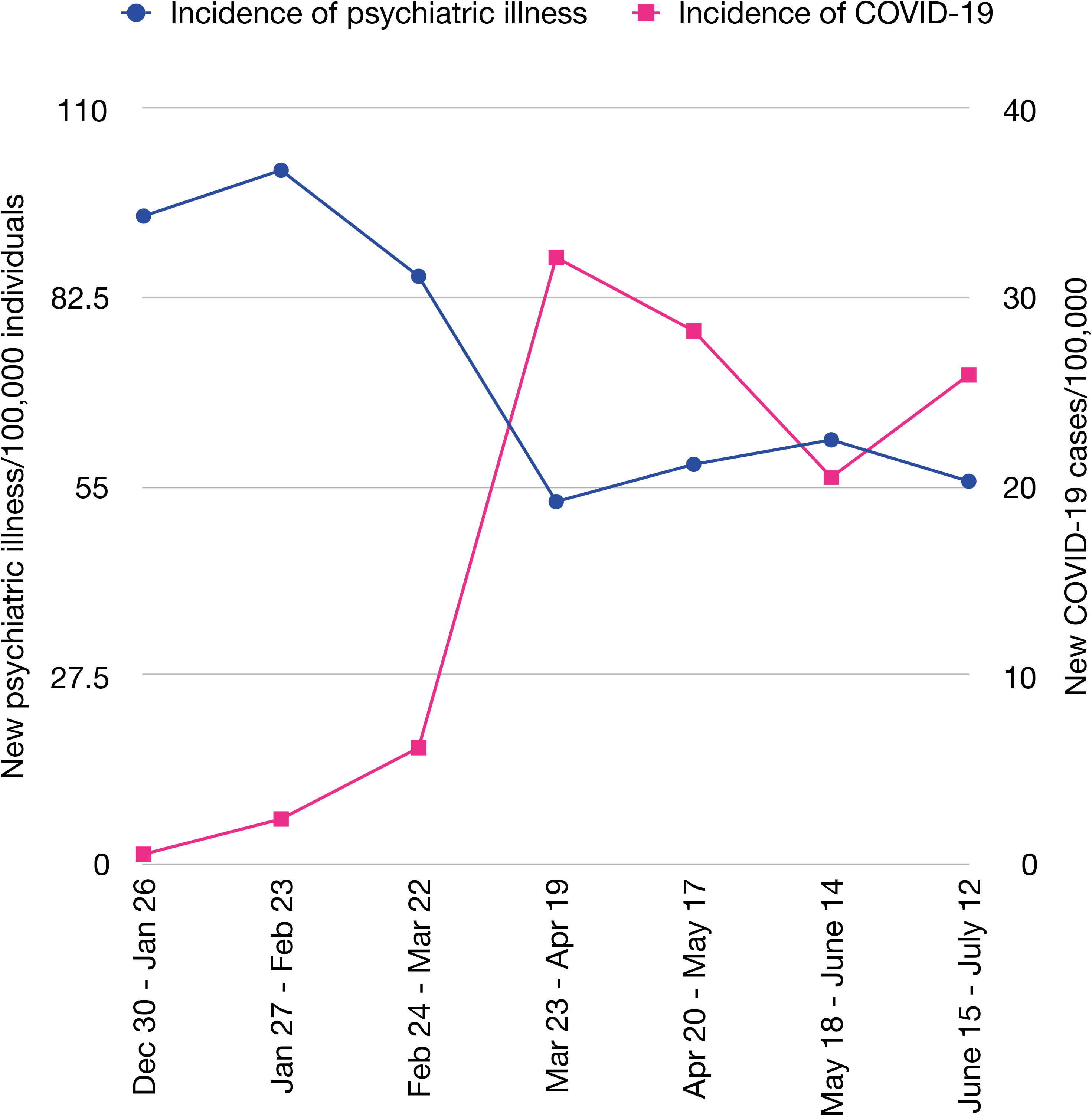
Incidence of first diagnoses of psychiatric illness and COVID-19 per 100,000 individuals per 4 weeks

Contextual factors provide part of the explanation for the difference in psychiatric sequelae between COVID-19 and control health events. Indeed all health events had significantly higher rates of psychiatric sequelae when they occurred after (vs. before) April 1, 2020 (HR comparing the period before to the period after April 1 ranging from 1.32 and 1.79, all p<0.05,, appendix p. 31) and the HR between COVID-19 and control health events were lower when these events occurred after April 1, 2020 (HR 1.31–1.83 vs. 1.58.2.24 when considering the whole study period, appendix p. 32).

However, these HR remained statistically larger than 1 indicating that contextual factors alone are insufficient to explain differences in psychiatric sequelae. In other words, experiencing *any* health event after (vs. before) April 1, 2020 led to a significantly higher rate of psychiatric sequelae but this rate was higher still after experiencing COVID-19.

### Psychiatric antecedents of COVID-19

Having a diagnosis of psychiatric illness in the year before the COVID-19 outbreak was associated with a 65% increased risk of COVID-19 (RR 1.65, 95% CI: 1.59–1.71, p<0.0001) compared to a cohort matched for established physical risk factors for COVID-19 (Fig. 3). The RR was higher in older patients (odds ratio 1.25, 95% CI: 1.14–1.38, p<0.0001).

**Fig. 3.**
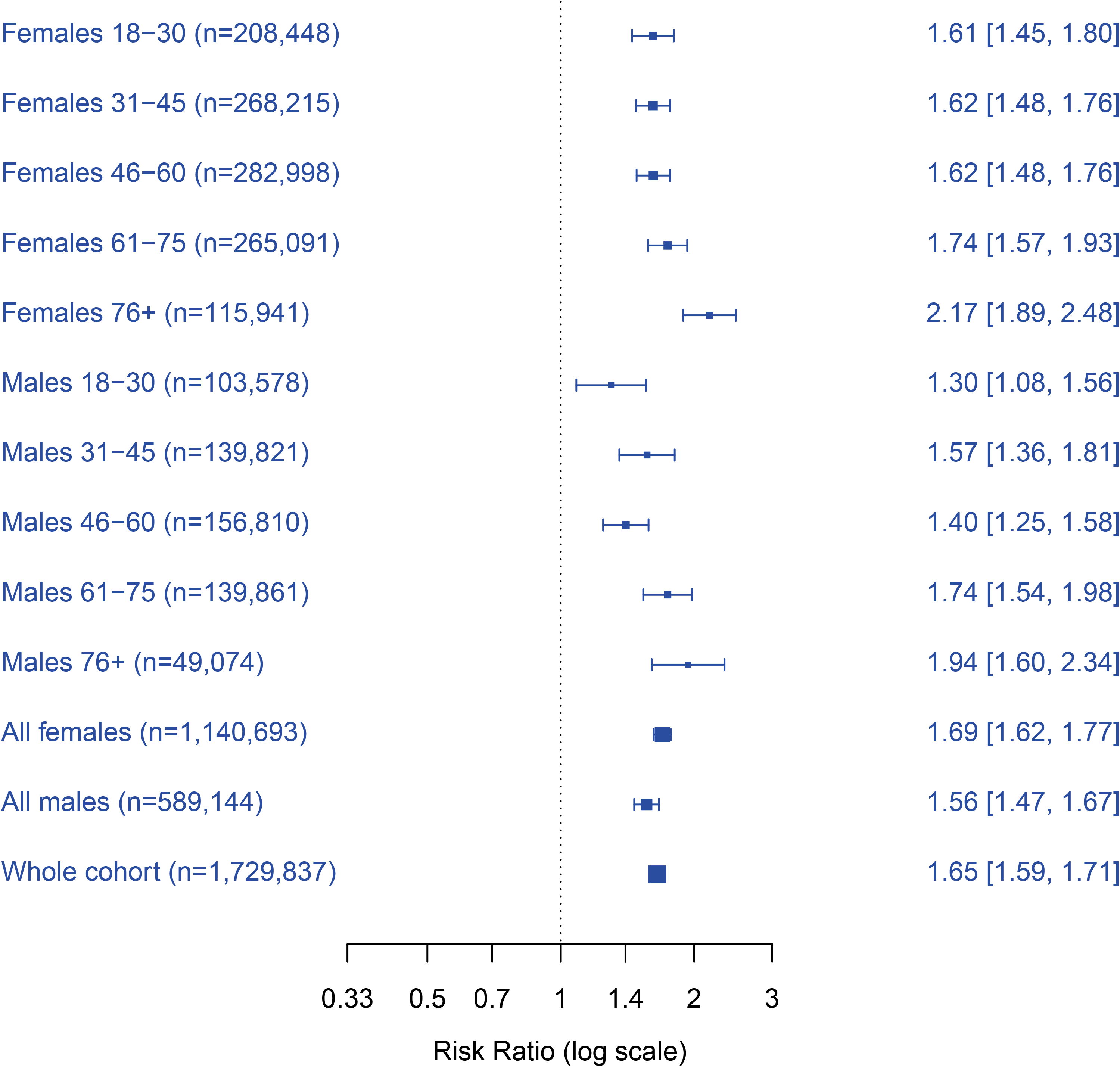
Relative risks of COVID-19 among patients with a psychiatric illness recorded in the past year compared to a matched cohort of patients with no history of psychiatric illness. Error bars and numbers in brackets represent 95% CI.

These results were robust to changes in the cohort specification, with a RR of 1.67 (95% CI: 1.57–.79, p< 0.0001, appendix p. 33) if this was a first psychiatric diagnosis, a RR of 1.80 (95% CI: 1.74–.86, p< 0.0001, appendix p. 33) among patients with a psychiatric diagnosis in the past three years, and a RR of 1.64 (95% CI: 1.58–1.70, p<0.0001, appendix p. 34) among patients whose race was known. Furthermore, a significantly elevated risk was also observed if the cohorts were limited to patients without any of the physical comorbidities that are risk factors for COVID-19 (RR: 1.64 (95% CI: 1.58–1.70, p< 0.57, 95% CI: 1.39–1.76, p< 0.0001, appendix p. 35). The finding was also robust if the outcome was limited to a confirmed diagnosis of COVID-19 (RR: 1.57, 95% CI: 1.51–1.64, p< 0.0001, appendix p. 36).

Only small differences in the RR of COVID-19 were observed when comparing classes of psychiatric diagnoses against each other: the RR among patients with a psychotic disorder were 1.17 (95% CI: 1. 02–1.33, p = 0.022) when compared a mood disorder and 1.08 (95% CI: 0.95–1.23, p = 0.22) when compared to an anxiety disorder. When compared to an anxiety disorder, the RR among patients with a mood disorder was 0.95 (95% CI: 0.92–0.99, p = 0.020).

## Discussion

We provide strong evidence that COVID-19 survivors have a significantly elevated rate of psychiatric disorders. We also show that a psychiatric history is associated with an increased risk of being diagnosed with COVID-19.

In the period between 14 days and 90 days after diagnosis, 5.8% of COVID-19 survivors had their first recorded diagnosis of psychiatric illness (F20-F48), compared to 2.5–3.4% of patients in the comparison cohorts. The figures are a minimum estimate, since there are likely to be many other patients who have not yet presented or received a diagnosis. Indeed, the Kaplan-Meier curves for COVID-19 show no signs of plateauing, suggesting that the absolute risk of psychiatric disorder will continue to increase with longer duration of follow-up. The HR from COVID-19 were higher compared to all other cohorts, indicating that COVID-19 has a specific impact on psychiatric health above and beyond changes that may be occurring in the general population. Evidence of the latter is apparent from the overall reduction in psychiatric diagnoses during the COVID-19 period, and by the fact that the risk of psychiatric sequelae remains higher for COVID-19 than for other health events in the ‘lockdown period’ (i.e. after April 1, 2020). The mechanism of COVID-19’s impact on mental health is unknown and requires urgent investigation. The modest relationship between severity of the illness (as proxied by inpatient admission) and psychiatric outcomes might represent a dose-response relationship suggesting that the association may at least partially be mediated by biological factors directly related to COVID-19 (e.g. viral load, immune response)^9,25,26^.

The psychiatric effects of COVID-19 were broad but not uniform. The HR was greater for anxiety disorders (ranging from 1 .6 –2.6) than for mood disorders (1.1–1.8). The particular impact of COVID-19 on anxiety is in line with expectations and highlights the need for effective and accessible interventions. The data show increased diagnoses in all major anxiety disorder categories, and so it remains unclear whether post-COVID-19 anxiety will have a particular PTSD-like picture. Rates of an insomnia diagnosis were also markedly elevated (HR 1.9–3.3), again in keeping with predictions that circadian disturbances will follow COVID-19 infection. In contrast, we did not find a clear signal for psychotic disorders despite case reports suggesting that this might occur^13,27^. Lastly, the 2–3 fold increased risk of dementia after COVID-19 extends findings from previous case series^13,28^ and is particularly concerning. Detailed investigation of this group should be a research priority, as should evaluation of other severe neuropsychiatric phenotypes which become apparent.

We had not anticipated that psychiatric history would be a risk factor for COVID-19, independent of known physical health factors. The finding appears robust, being seen in all age strata, and in both sexes, and is substantial—a 1.65-fold excess. The basis for the finding is unclear. It is not related to any specific diagnostic category, and it was similar regardless of whether the diagnosis was made within one or three years, and whether or not the known physical risk factors for COVID-19 were present. These features suggest that broad or non-specific factors related to psychiatric illness are involved, potentially including cognitive impairment, lower awareness of risk, or increased difficulty in following social distancing recommendations^4^. Residual confounding, for example by unmeasured lifestyle factors, is another possible explanation. It may also be that vulnerability to COVID-19 is increased by the pro-inflammatory state postulated to occur in some forms of psychiatric disorder, or be related to psychotropic medication.

The strengths of this study are the sample size, the amount of data available, the use of propensity score matching to control for confounding, the range of sensitivity analyses, and the real-world nature of the data. The study also has limitations. Despite the matching and use of various comparison cohorts, there may be residual confounding. There is little information in the network regarding social and economic factors which might influence clinical outcomes post COVID-19. We do not know whether psychiatric diagnoses were made correctly and consistently between cohorts; it is possible that clinicians were more likely to diagnose a psychiatric illness after a COVID-19 diagnosis due to subjective bias. We did not examine whether COVID-19 affects relapse rate in those with prior psychiatric disorder. The results cannot necessarily be generalised to other populations or healthcare settings.

Nevertheless, the findings are of sufficient robustness and magnitude to have some immediate implications. The figures provide minimum estimates of the excess in psychiatric morbidity to be anticipated in survivors of COVID-19 and for which services need to plan^29^. As COVID-19 sample sizes and survival times increase, it will be possible not only to refine these findings, but also to identify rarer and delayed psychiatric presentations. It will also be important to explore additional risk factors for contracting COVID-19, and for developing psychiatric illness thereafter, since some elements may prove to be modifiable.

## Data Availability

Data is only available via the TriNetX platform to approved users.

https://live.trinetx.com

## Research in context

### Evidence before this study

From January 1 to August 1, 2020, we searched PubMed with the terms: (COVID-19 OR SARS-CoV2 OR SARS-CoV-2) AND (pysch* OR cognit* OR mental) and MedRxiv with the terms COVID-19 OR SARS-CoV2 OR SARS-CoV-2 in the ‘neurology’ and/or ‘psychiatry’ categories. We also manually reviewed the references in the identified papers. Studies investigating the psychiatric consequences of COVID-19 lacked a control condition, consisted mostly of surveys, and mostly used self-reported symptoms (rather than diagnoses) as an outcome. No study has assessed the risk of developing psychiatric sequelae over time and only anecdotal evidence exists for the risk of dementia as a potential consequence of COVID-19. In terms of psychiatric risk factors for COVID-19, two case-control studies were identified. One study investigated risk factors for hospitalisation for (rather than diagnosis of) COVID-19. The other study used historical data (not acquired during the same period as COVID-19) as a control group. As these were case-control studies, only odds-ratios could be estimated rather than relative or absolute risks. In addition, in both studies, controls were not well matched to cases.

### Added value of this study

This is the first dataset allowing the psychiatric sequelae and antecedents of COVID-19 to be measured. The study cohorts are orders of magnitude larger than previous studies producing more precise, more representative estimates of even small but important effects—such as the incidence of dementia. It uses propensity score matching to control for many variables, including established physical risk factors for COVID-19 and for more severe COVID-19 illness, and it uses large-scale real-world data thus providing more clinically relevant findings. We used time-to-event data for the analysis of psychiatric sequelae, thus providing the first evidence for their temporal evolution. Our findings show that COVID-19 survivors have significantly higher rates of psychiatric diagnoses (in particular, anxiety disorders, but also depression, insomnia, and dementia) and also show that a psychiatric history is a risk factor for being diagnosed with COVID-19 independent of known physical risk factors.

### Implications of all the available evidence

The implications of the available evidence are three-fold. First, adequate service provision should be made available to face the increased incidence of psychiatric illness following COVID-19. Second, psychiatric follow up should be considered for patients who survived COVID-19. Third, past psychiatric history should be routinely queried during the anamnesis of a patient presenting with COVID-19 symptoms to adjust pre-test probability.

## Acknowledgements

MT is a National Institute for Health Research (NIHR) Academic Clinical Fellow. JRG and PJH are supported by the NIHR Oxford Health Biomedical Research Centre (grant BRC-1215–20005). The views expressed are those of the authors and not necessarily those of the National Health Service, NIHR, or the Department of Health.

## Author contributions

PJH and MT designed the study. MT carried out data analyses. SL and JRG assisted with data analysis and interpretation. MT and PJH wrote the paper with input from JRG and SL. PJH is the guarantor.

## References

1 Xiang Y-T, Yang Y, Li W, et al. Timely mental health care for the 2019 novel coronavirus outbreak is urgently needed. Lancet Psychiatry 2020; 7: 228–9.

2 World Health Organization. Mental health and psychosocial considerations during the COVID-19 outbreak, 18 March 2020. World Health Organization, 2020 https://apps.who.int/iris/bitstream/handle/10665/331490/WHO-2019-nCoV-MentalHealth-2020.1-eng.pdf.

3 Holmes EA, O’Connor RC, Perry VH, et al. Multidisciplinary research priorities for the COVID-19 pandemic: a call for action for mental health science. Lancet Psychiatry 2020; 7; 547–60. DOI:10.1016/S2215-0366(20)30168-1.

4 Yao H, Chen J-H, Xu Y-F. Patients with mental health disorders in the COVID-19 epidemic. Lancet Psychiatry 2020; 7: e21.

5 Hu Y, Chen Y, Zheng Y, et al. Factors related to mental health of inpatients with COVID-19 in Wuhan, China. Brain Behav Immun 2020; published online July 15. DOI:10.1016/j.bbi.2020.07.016.

6 Nie X-D, Wang Q, Wang M-N, et al. Anxiety and depression and its correlates in patients with coronavirus disease 2019 in Wuhan. Int J Psychiatry Clin Pract 2020;: 1–6.

7 Speth MM, Singer Cornelius T, Oberle M, Gengler I, Brockmeier SJ, Sedaghat AR. Mood, anxiety and olfactory dysfunction in COVID 19: evidence of central nervous system involvement? Laryngoscope 2020; published online July 2. DOI:10.1002/lary.28964.

8 Paz C, Mascialino G, Adana-Díaz L, et al. Anxiety and depression in patients with confirmed and suspected COVID-19 in Ecuador. Psychiatry Clin Neurosci 2020; published online July 1. DOI:10.1111/pcn.13106.

9 Mazza MG, De Lorenzo R, Conte C, et al. Anxiety and depression in COVID-19 survivors: role of inflammatory and clinical predictors. Brain Behav Immun 2020; published online July 29. DOI:10.1016/j.bbi.2020.07.037.

10 Halpin SJ, McIvor C, Whyatt G, et al. Post discharge symptoms and rehabilitation needs in survivors of COVID 19 infection: a cross sectional evaluation. J Med Virol 2020; published online July 30. DOI:10.1002/jmv.26368.

11 Ma Y-F, Li W, Deng H-B, et al. Prevalence of depression and its association with quality of life in clinically stable patients with COVID-19. J Affect Disord 2020; 275: 145–8.

12 Zhang J, Lu H, Zeng H, et al. The differential psychological distress of populations affected by the COVID-19 pandemic. Brain Behav Immun 2020; 87: 49–50.

13 Varatharaj A, Thomas N, Ellul MA, et al. Neurological and neuropsychiatric complications of COVID-19 in 153 patients: a UK-wide surveillance study. Lancet Neurology published online June 25, 2020. DOI:https://doi.org/10.1016/S2215-0366(20)30287-X

14 Rogers JP, Chesney E, Oliver D, et al. Psychiatric and neuropsychiatric presentations associated with severe coronavirus infections: a systematic review and meta-analysis with comparison to the COVID-19 pandemic. Lancet Psychiatry 2020; 7: 611–27.

15 Batty GD, Deary IJ, Luciano M, Altschul DM, Kivimäki M, Gale CR. Psychosocial factors and 17/22 hospitalisations for COVID-19: Prospective cohort study based on a community sample. Brain, Behavior, and Immunity On line Jun 17, 2020. doi: 10.1016/j.bbi.2020.06.021 [Epub ahead of print]

16 Chang TS, Ding Y, Freund MK, et al. Prior diagnoses and medications as risk factors for COVID-19 in a Los Angeles Health System. medRxiv 2020; published online July 4. DOI:10.1101/2020.07.03.20145581.

17 Harrison PJ, Luciano S, Colbourne L. Rates of delirium associated with calcium channel blockers compared to diuretics, renin-angiotensin system agents and beta-blockers: An electronic health records network study. J Psychopharmacol 2020; 34: 848–55.

18 de Lusignan S, Dorward J, Correa A, et al. Risk factors for SARS-CoV-2 among patients in the Oxford Royal College of General Practitioners Research and Surveillance Centre primary care network: a cross-sectional study. Lancet Infect Dis 2020; published online May 15. DOI:10.1016/S1473-3099(20)30371-6.

19 Zhang J-J, Dong X, Cao Y-Y, et al. Clinical characteristics of 140 patients infected with SARSCoV-2 in Wuhan, China. Allergy 2020. https://onlinelibrary.wiley.com/doi/abs/10.1111/all.14238.

20 Chen N, Zhou M, Dong X, et al. Epidemiological and clinical characteristics of 99 cases of 2019 novel coronavirus pneumonia in Wuhan, China: a descriptive study. Lancet 2020; 395: 507–13.

21 Williamson EJ, Walker AJ, Bhaskaran K, et al. OpenSAFELY: factors associated with COVID-19 death in 17 million patients. Nature 2020; published online July 8. DOI:10.1038/s41586-020-2521-4.

22 Austin PC. An introduction to propensity score methods for reducing the effects of confounding in observational studies. Multivariate Behav Res 2011; 46: 399–424.

23 Haukoos JS, Lewis RJ. The propensity score. JAMA 2015; 314: 1637–8.

24 Grambsch PM, Therneau TM. Proportional hazards tests and diagnostics based on weighted residuals. Biometrika 1994; 81: 515–26.

25 Postolache TT, Benros ME, Brenner LA. Targetable biological mechanisms implicated in emergent psychiatric conditions associated with SARS-CoV-2 infection. JAMA Psychiatry 2020; published online July 31. DOI:10.1001/jamapsychiatry.2020.2795.

26 Gupta A, Madhavan MV, Sehgal K, et al. Extrapulmonary manifestations of COVID-19. Nat Med 2020; 26: 1017–32.

27 Parra A, Juanes A, Losada CP, et al. Psychotic symptoms in COVID-19 patients. A retrospective descriptive study. Psychiatry Res 2020; 291: 113254.

28 Pinna P, Grewal P, Hall JP, et al. Neurological manifestations and COVID-19: Experiences from a tertiary care center at the Frontline. J Neurol Sci 2020; 415: 116969.

29 Greenhalgh T, Knight M, A’Court C, Buxton M, Husain L. Management of post-acute covid-19 in primary care. BMJ 2020; 370: m3026.

